# Association Between Systemic Inflammation Response Index (SIRI) and Cardiovascular Disease Mortality in Individuals with Anemia: Findings from NHANES 1999–2018

**DOI:** 10.1101/2025.10.21.25338494

**Authors:** Xiangkuan Cheng, Lanling Liu, Jie Wu, Yueming Tian, Xiuluan Du, Zhibin Li, Yuansheng Lin

## Abstract

**Background:** Cardiovascular disease (CVD) is a leading cause of mortality in individuals with anemia. This study investigates the association between the Systemic Inflammation Response Index (SIRI) and CVD mortality in this population.

**Methods:** Data from 3,212 participants with confirmed anemia from the NHANES 1999–2018 were analyzed. Participants were stratified by SIRI levels. Univariate and multivariate Cox regression analyses assessed the relationship between SIRI and CVD mortality, with subgroup analyses for various demographic and clinical factors.

**Results:** Higher SIRI levels were associated with older age, male gender, and increased prevalence of chronic conditions, such as chronic kidney disease, diabetes, and hypertension. CVD mortality was significantly higher in the highest SIRI group (15.7%) compared to the lowest (4.9%, p < 0.001). SIRI was a significant predictor of CVD mortality (HR: 1.3, 95% CI: 1.25–1.36, p < 0.001). Participants in the highest SIRI quartile had a markedly increased risk (HR: 4.24, 95% CI: 2.96–6.07, p < 0.001). A non-linear relationship was observed with a threshold at SIRI 0.244, above which the risk steeply increased (HR: 4.807, 95% CI: 2.945–7.845, p < 0.001).

**Conclusions:** Higher SIRI levels are strongly associated with increased CVD mortality in individuals with anemia, with a non-linear relationship. These findings highlight the role of systemic inflammation in CVD risk, suggesting that SIRI may be a valuable biomarker in this population.

## Introduction

Cardiovascular disease (CVD) remains one of the leading causes of mortality globally, with a particularly high burden among individuals with chronic conditions such as anemia (1, 2). Anemia, characterized by a deficiency in the number or quality of red blood cells, has been shown to increase the risk of adverse cardiovascular outcomes, including heart failure and CVD mortality (3, 4). Recent evidence suggests that systemic inflammation may play a key role in exacerbating this risk (5, 6). The Systemic Inflammation Response Index (SIRI), a composite marker of inflammation that incorporates white blood cell count and its subtypes, has emerged as a potential biomarker for assessing inflammation and predicting cardiovascular outcomes in various populations (7).

While the relationship between inflammation and CVD is well established in general populations, its role in individuals with anemia has not been extensively studied. In this context, SIRI has been proposed as a promising indicator of inflammation that may help identify individuals at higher risk for CVD mortality. Previous research has highlighted the significance of inflammation in cardiovascular health, but few studies have examined how this marker interacts with anemia in predicting CVD outcomes (8). Additionally, the potential non-linear nature of this association, along with its varying impact across different subgroups, has not been thoroughly explored.

This study aims to investigate the relationship between SIRI and CVD mortality in individuals with anemia using data from the National Health and Nutrition Examination Survey (NHANES) 1999–2018. We hypothesize that higher SIRI levels are associated with an increased risk of CVD mortality in the anemic population, and that this association may vary by demographic and clinical factors. By examining baseline characteristics, performing univariate and multivariate regression analyses, and exploring non-linear relationships, we aim to provide a comprehensive understanding of how systemic inflammation, as measured by SIRI, influences cardiovascular risk in this vulnerable group. This study’s findings could inform targeted strategies for the prevention and management of CVD in individuals with anemia.

## Methods

### Study Population

Data for this study were obtained from the National Health and Nutrition Examination Survey (NHANES) 1999–2018. Initially, 55,081 participants aged 20 years and older were included. Pregnant participants (n=1,491) and those with missing data on key variables (n=15,765) were excluded. After these exclusions, a final cohort of 37,925 participants remained. Of these, 34,713 participants without confirmed anemia were further excluded, leaving 3,212 participants with confirmed anemia for analysis (Figure 1). Ethical approval for the research protocol and associated consent documentation (Protocol #98-12) was obtained through the NHANES Institutional Review Board’s Ethics Committee. This study did not prospectively recruit human participants (e.g., no clinical trials, questionnaires, or new data/sample collection). It is a retrospective analysis of pre-existing, fully anonymized data from the NHANES database. Therefore, no additional recruitment period or consent procedures were required.

**Figure 1.**
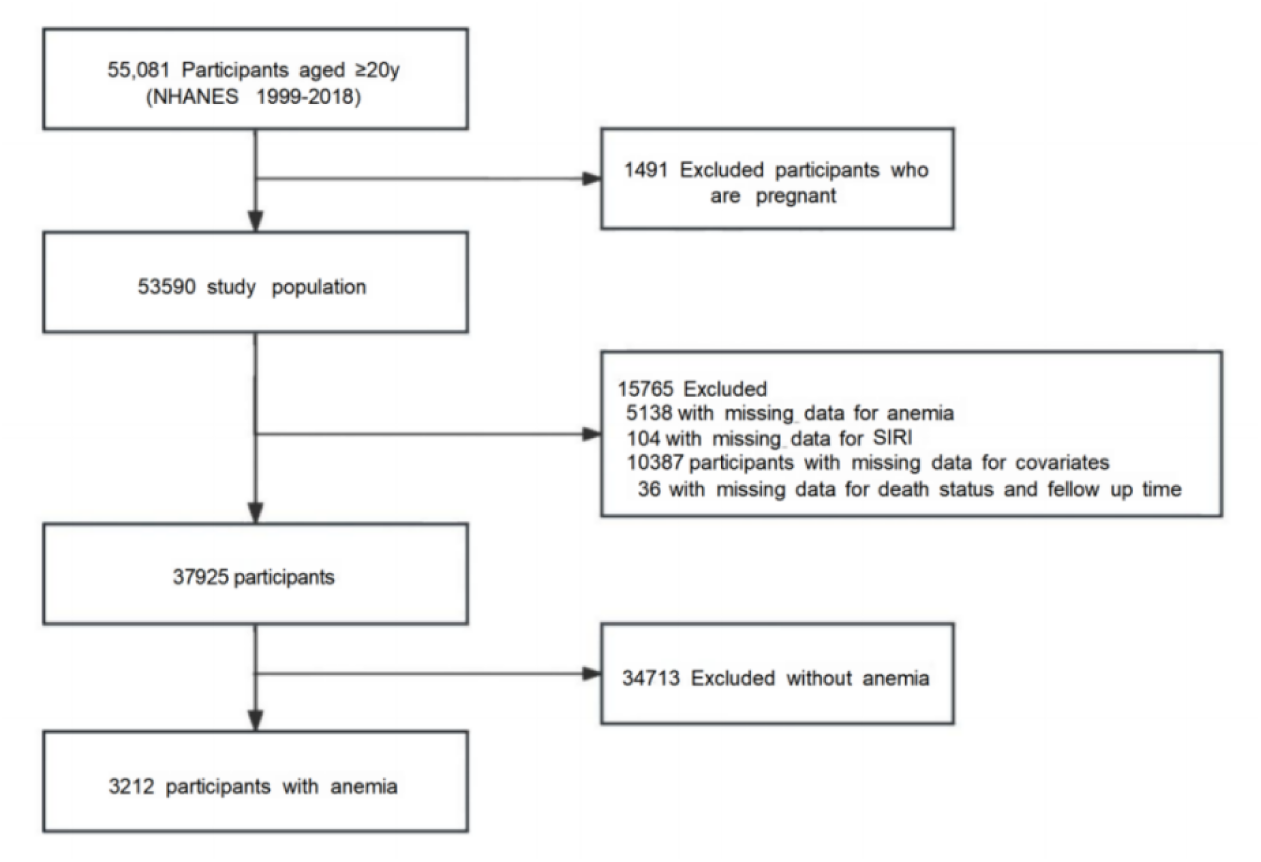
Flowchart of patient selection

### SIRI Calculation

The SIRI (9)was calculated using the following formula: SIRI = (Neutrophil count × Monocyte count) / Lymphocyte count

Participants were stratified into quartiles based on their SIRI levels, with the lowest quartile (<0.67) representing participants with the least systemic inflammation and the highest quartile (≥ 1.65) representing participants with the most inflammation.

### Baseline Characteristics

Baseline characteristics of the study participants, including age, gender, race/ethnicity, education level, and health conditions (chronic kidney disease, diabetes, hypertension, stroke, coronary heart disease), were compared across SIRI groups. Continuous variables were summarized as means (SD) or medians (IQR), and categorical variables were summarized using frequencies and percentages. Comparisons between groups were made using analysis of variance (ANOVA) for continuous variables and chi-square tests for categorical variables.

### Univariate Cox Regression Analysis

Univariate Cox regression analysis (10) was performed to identify potential predictors of CVD mortality among participants with anemia. The primary independent variable of interest was SIRI, with additional adjustments for demographic variables (age, gender, ethnicity), health conditions (chronic kidney disease, diabetes, hypertension, stroke, coronary heart disease), and lifestyle factors (smoking and alcohol consumption). Hazard ratios (HRs) with 95% confidence intervals (CIs) were calculated for each variable.

### Multivariate Cox Regression Analysis

To adjust for potential confounders, multivariate Cox regression analysis (11) was conducted. Models were adjusted for age, gender, race/ethnicity, educational attainment, health conditions, and lifestyle factors. Three models were tested: Model 1 adjusted for basic demographics and health conditions. Model 2 included additional adjustment for lifestyle factors (smoking, alcohol use). Model 3 included all potential confounders. Hazard ratios (HRs) and 95% confidence intervals (CIs) were calculated for each model.

### Non-linear Association Between SIRI and CVD Mortality

To explore potential non-linearity in the relationship between SIRI and CVD mortality, we used spline regression analysis and identified a significant threshold in the SIRI distribution. A likelihood ratio test was conducted to compare the non-linear model with a linear model. Two distinct slopes were examined based on the identified threshold of SIRI = 0.244.

### Subgroup Analyses

Subgroup analyses (12) were conducted to assess the consistency of the association between SIRI and CVD mortality across different demographic and clinical factors, including gender, age, body mass index (BMI), chronic kidney disease, diabetes, stroke, cancer, and thyroid disease. Interaction terms were tested in multivariate models to assess potential effect modification.

### Statistical Analysis

All statistical analyses were performed with the statistical software packages R (http://www.R-project.org, The R Foundation) and Free Statistics software versions 2.0. A p-value of < 0.05 was considered statistically significant for all tests. Sampling weights provided by NHANES were applied to adjust for the complex survey design.

## Results

### Baseline Characteristics and Cardiovascular Disease Mortality in Participants with Anemia

In the NHANES (1999–2018), 55,081 participants aged ≥20 years were initially included. After excluding pregnant participants (n=1,491) and those with missing data (n=15,765), 37,925 participants remained. Of these, 34,713 without anemia were excluded, leaving 3,212 participants with confirmed anemia for analysis (Figure 1).

Significant differences in baseline characteristics were observed across groups stratified by SIRI levels (Table 1). Age increased with higher SIRI levels, from 50.8 years in the lowest SIRI group (<0.67) to 63.2 years in the highest SIRI group (≥ 1.65, p < 0.001). Gender distribution varied significantly, with more males in the highest SIRI group (51.4%, p < 0.001). Race/ethnicity differences were also significant, with Non-Hispanic White participants being more prevalent in the highest SIRI group (49.3%, p < 0.001). Educational attainment was lower in the lowest SIRI group (p = 0.038).

**Table 1.**
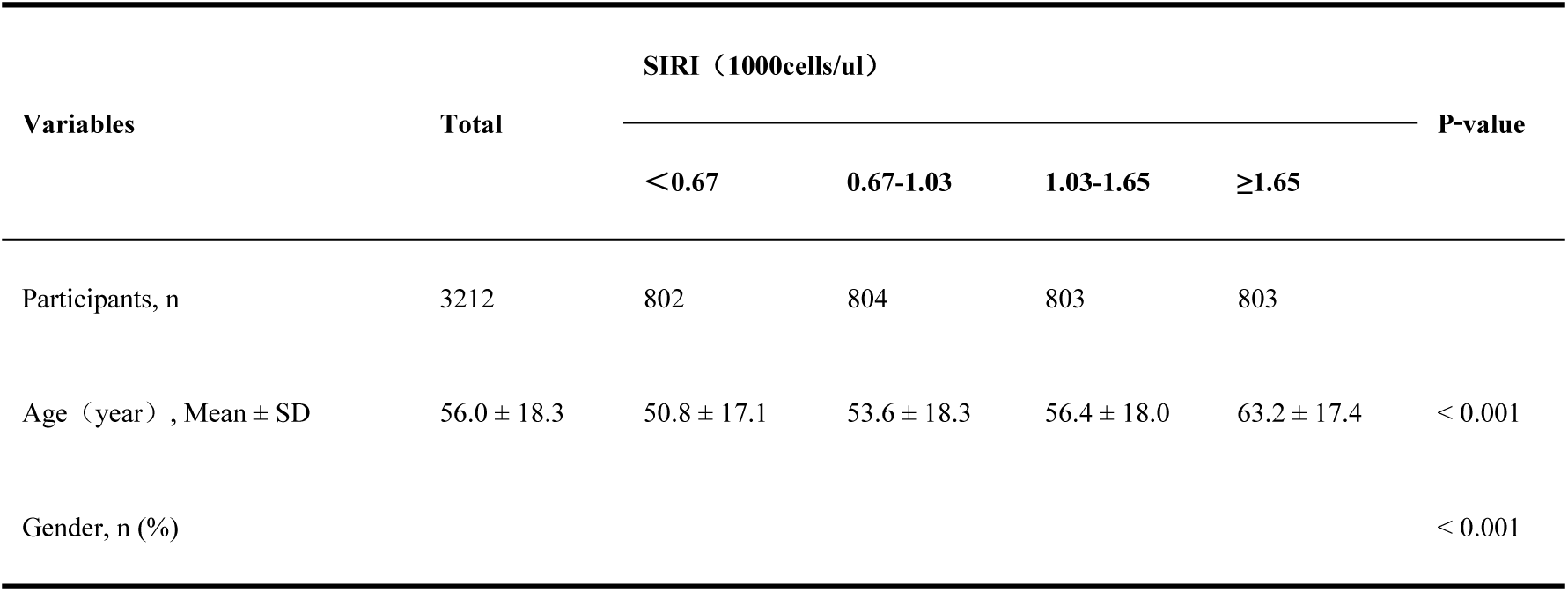
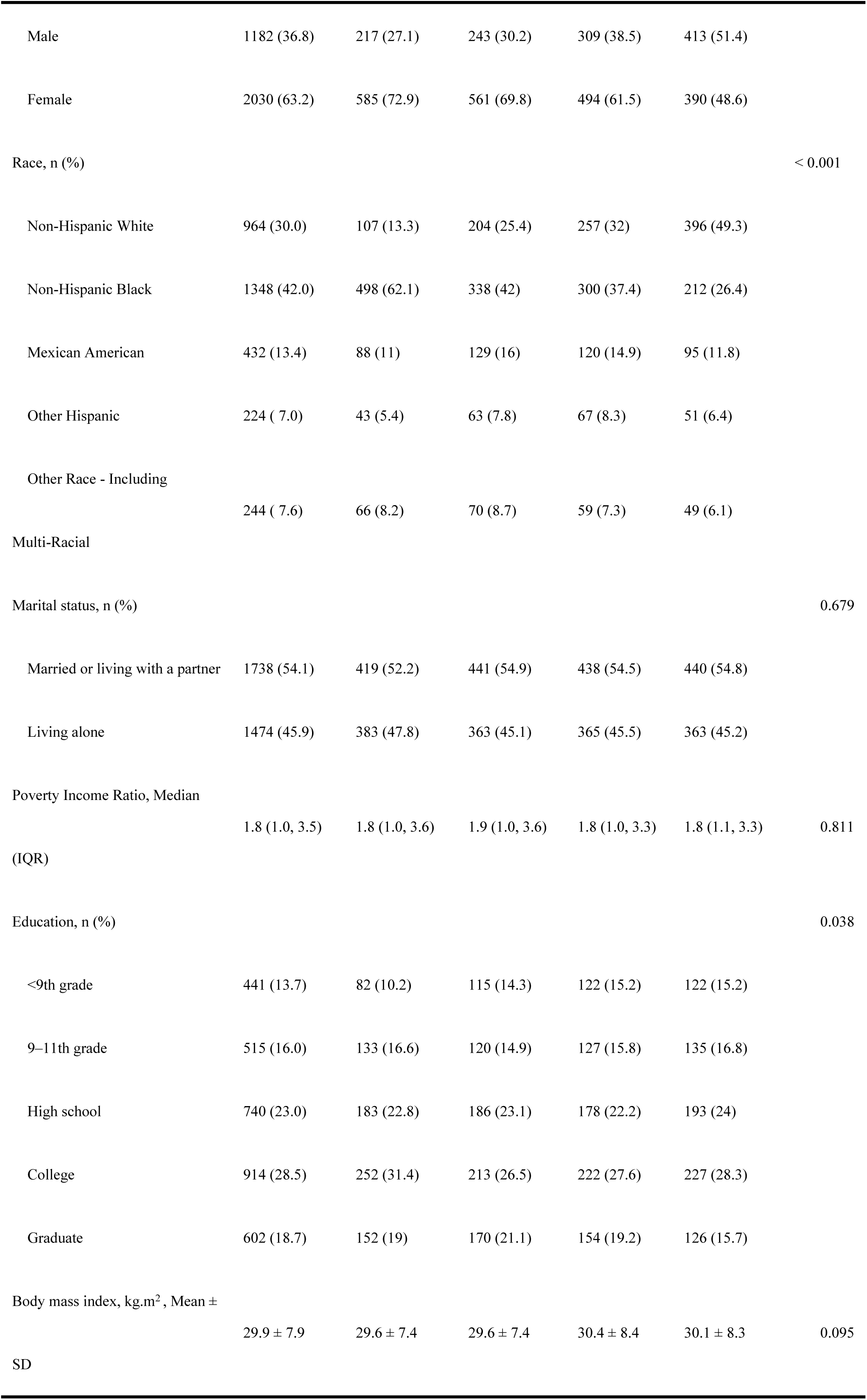
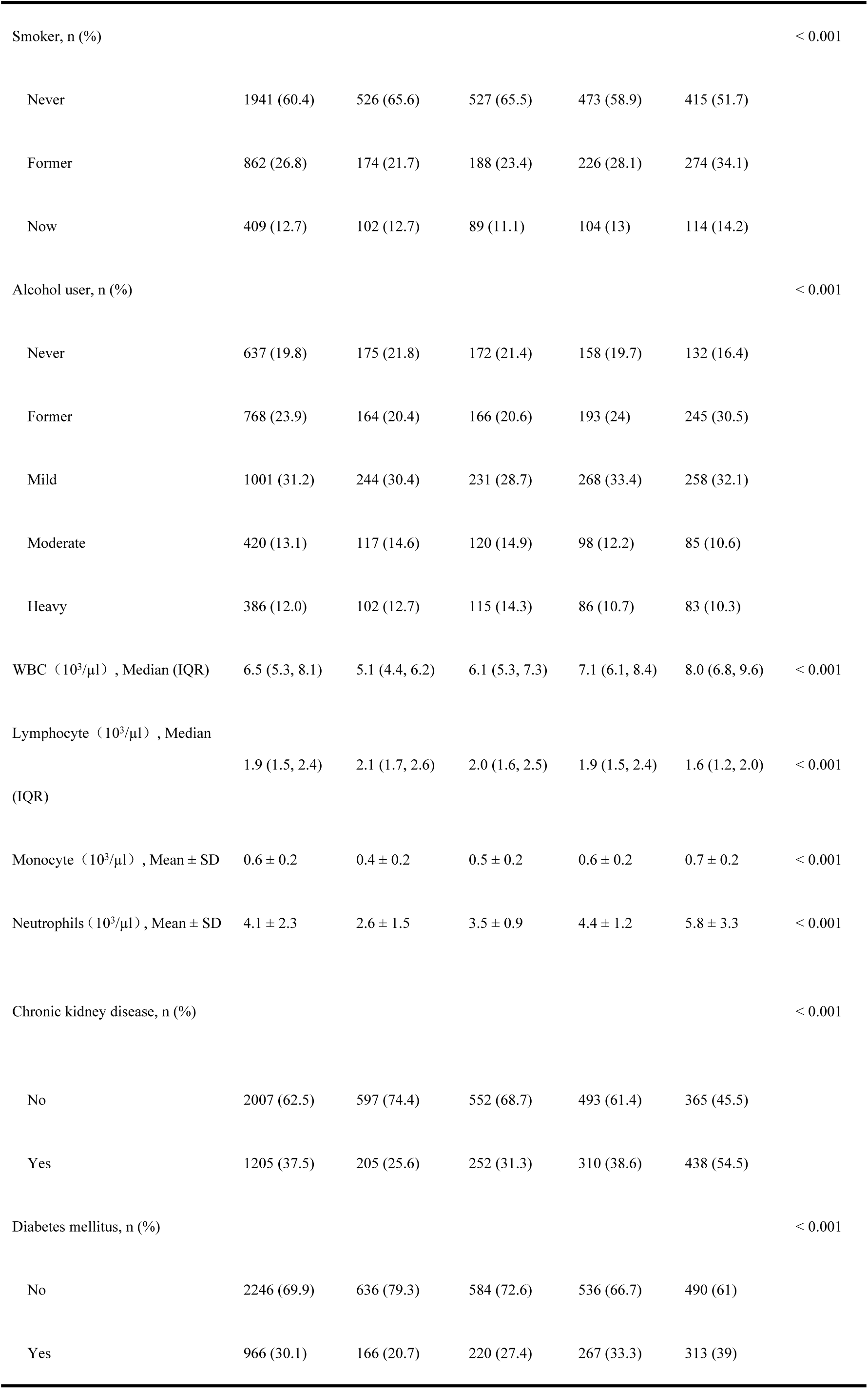
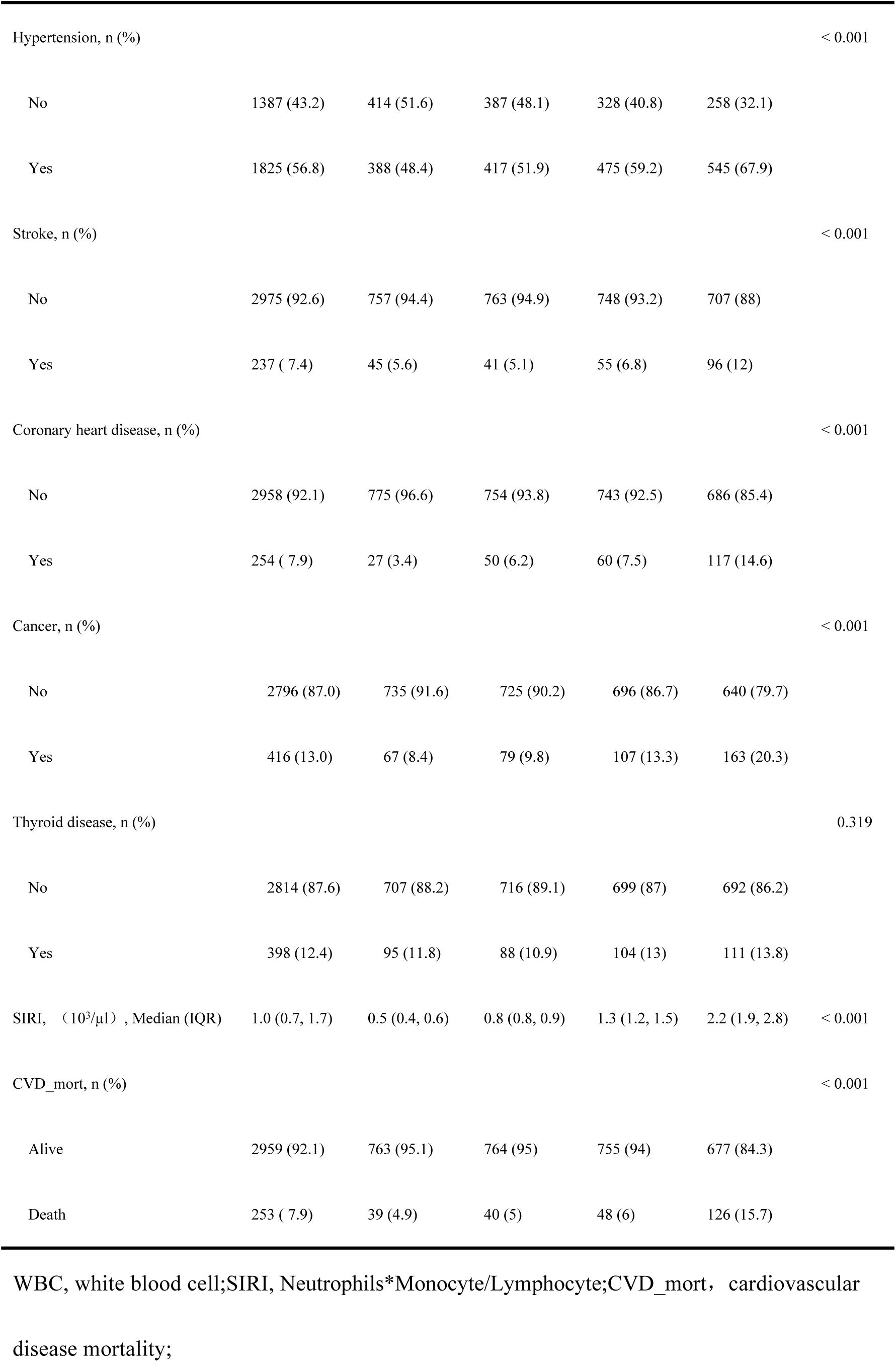
Basic characteristics of NHANES participants (N = 3212).

Higher SIRI levels were associated with an increased prevalence of chronic conditions, including chronic kidney disease (CKD), diabetes, hypertension, stroke, and coronary heart disease (CHD) (all p-values < 0.001). Specifically, 54.5% of participants in the highest SIRI group had CKD, compared to 25.6% in the lowest SIRI group. SIRI was also positively associated with white blood cell count, lymphocyte, monocyte, and neutrophil counts (all p-values < 0.001). The median SIRI level increased from 0.5 (0.4, 0.6) in the lowest group to 2.2 (1.9, 2.8) in the highest group (p < 0.001).

CVD mortality was higher in the highest SIRI group, with 15.7% of participants having died from CVD, compared to 4.9% in the lowest SIRI group (p < 0.001).

### Univariate Cox Regression Analysis of Predictors of CVD Mortality in Anemic Individuals

Univariate Cox regression analysis identified several significant predictors of CVD mortality in individuals with anemia (Table 2). Age was positively associated with CVD mortality (HR: 1.11, 95% CI: 1.09–1.12, p < 0.001). Female participants had a significantly lower risk of CVD mortality compared to males (HR: 0.29, 95% CI: 0.23–0.38, p < 0.001). Other significant predictors included ethnicity, education level, poverty income ratio, marital status, and smoking status. Health-related factors such as CKD (HR: 7.53, 95% CI: 5.60–10.11, p < 0.001), diabetes (HR: 2.54, 95% CI: 1.99–3.26, p < 0.001), hypertension (HR: 4.58, 95% CI: 3.30–6.34, p < 0.001), stroke (HR: 3.26, 95% CI: 2.32–4.59, p < 0.001), and CHD (HR: 4.63, 95% CI: 3.46–6.21, p < 0.001) were strongly associated with increased CVD mortality. Additionally, current smoking and heavy alcohol consumption were significant predictors (p < 0.001).

**Table 2.**
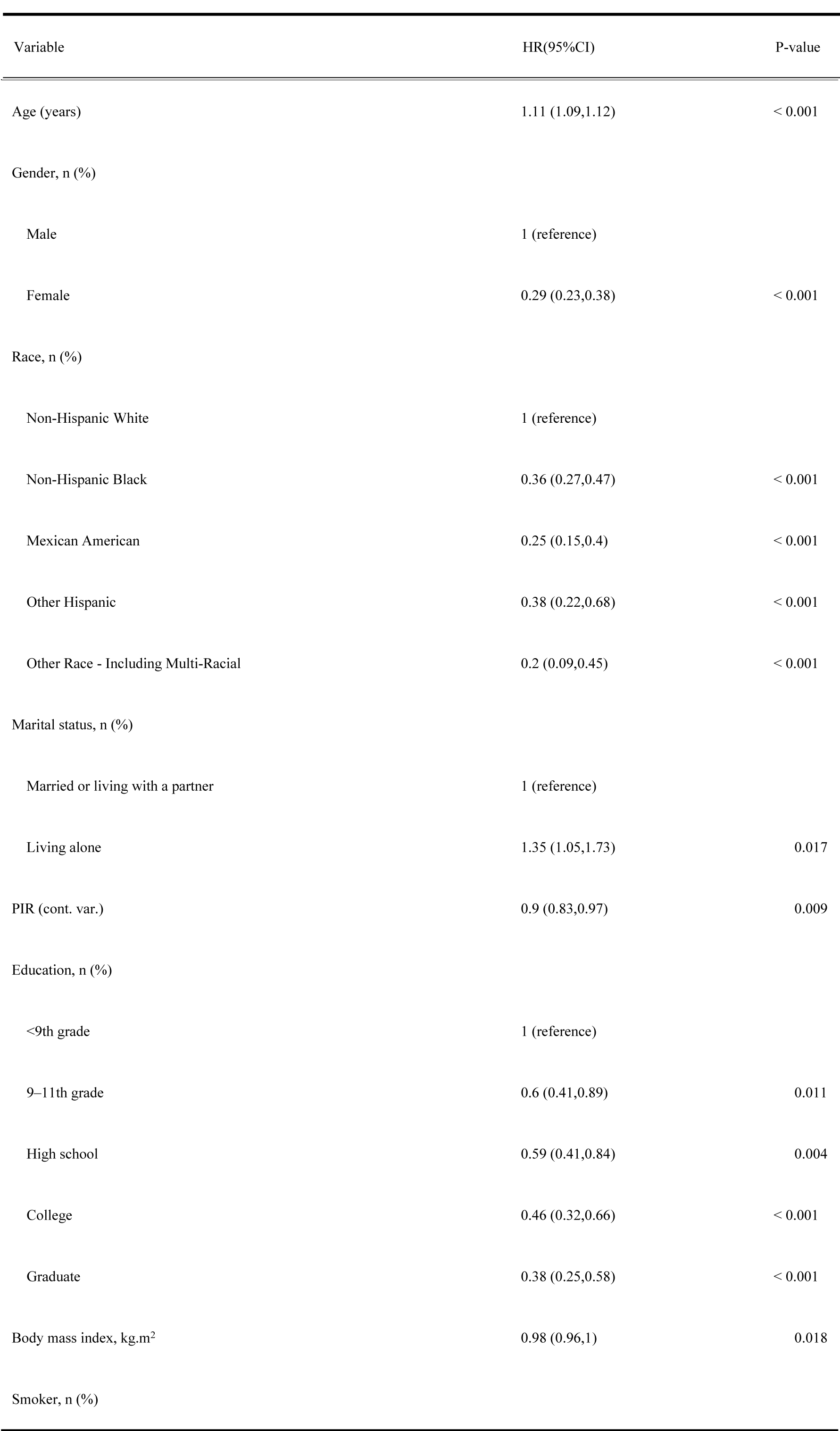
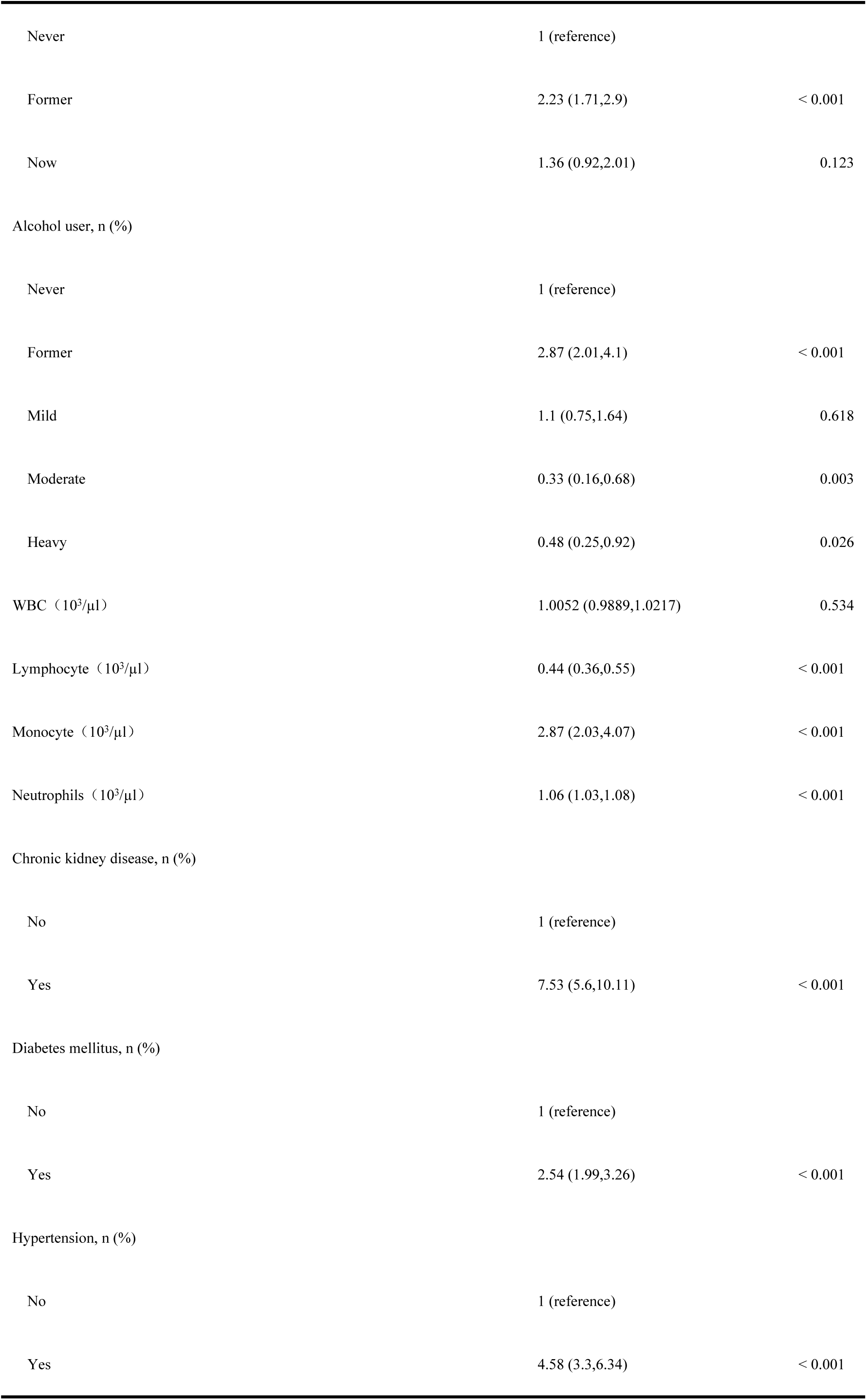
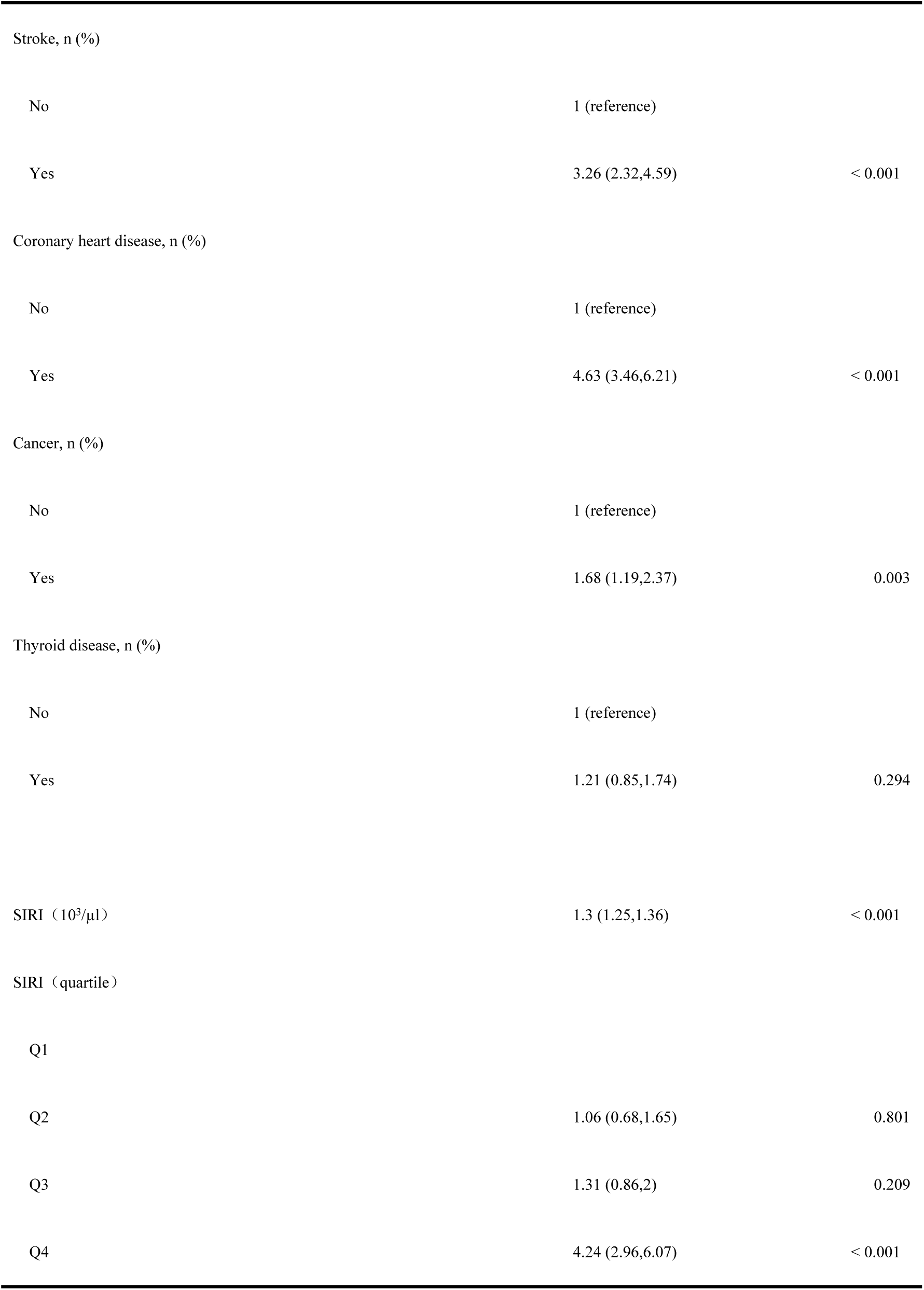
Univariate Cox Regression Analysis of Predictors of CVD Mortality in Anemic Individuals.

Regarding systemic inflammation, SIRI (HR: 1.3, 95% CI: 1.25–1.36, p < 0.001) was a significant continuous predictor of CVD mortality. Among the SIRI quartiles, participants in the highest quartile (≥1.65) had a markedly increased risk compared to the lowest quartile (HR: 4.24, 95% CI: 2.96–6.07, p < 0.001). Furthermore, monocyte (HR: 2.87, 95% CI: 2.03–4.07, p < 0.001) and neutrophil counts (HR: 1.06, 95% CI: 1.03–1.08, p < 0.001) were significantly associated with increased CVD mortality, while lymphocyte count was inversely associated (HR: 0.44, 95% CI: 0.36–0.55, p < 0.001).

### Multivariate Cox Regression Analysis of the Association Between SIRI and CVD Mortality

Multivariate Cox regression analysis further confirmed the significant association between SIRI and CVD mortality in individuals with anemia (Table 3). In the crude model, SIRI was significantly associated with increased CVD mortality (HR: 2.67, 95% CI: 2.23–3.20, p < 0.001). This association remained significant after adjusting for potential confounders in all models (HR in Model 1: 1.68, 95% CI: 1.36–2.09, p < 0.001; Model 2: 1.56, 95% CI: 1.25–1.94, p < 0.001; Model 3: 1.78, 95% CI: 1.38–2.30, p < 0.001).

Stratified by SIRI quartiles, participants in the highest quartile (Q4) consistently demonstrated a significantly higher risk of CVD mortality compared to those in the reference group (Q3). In the crude model, the HR for Q4 was 3.23 (95% CI: 2.32–4.51, p < 0.001), and it remained significant after adjusting for covariates in all models (HR in Model 1: 2.31, 95% CI: 1.65–3.24, p < 0.001; Model 2: 2.26, 95% CI: 1.61–3.17, p < 0.001; Model 3: 2.40, 95% CI: 1.69–3.40, p < 0.001).

**Table 3.**
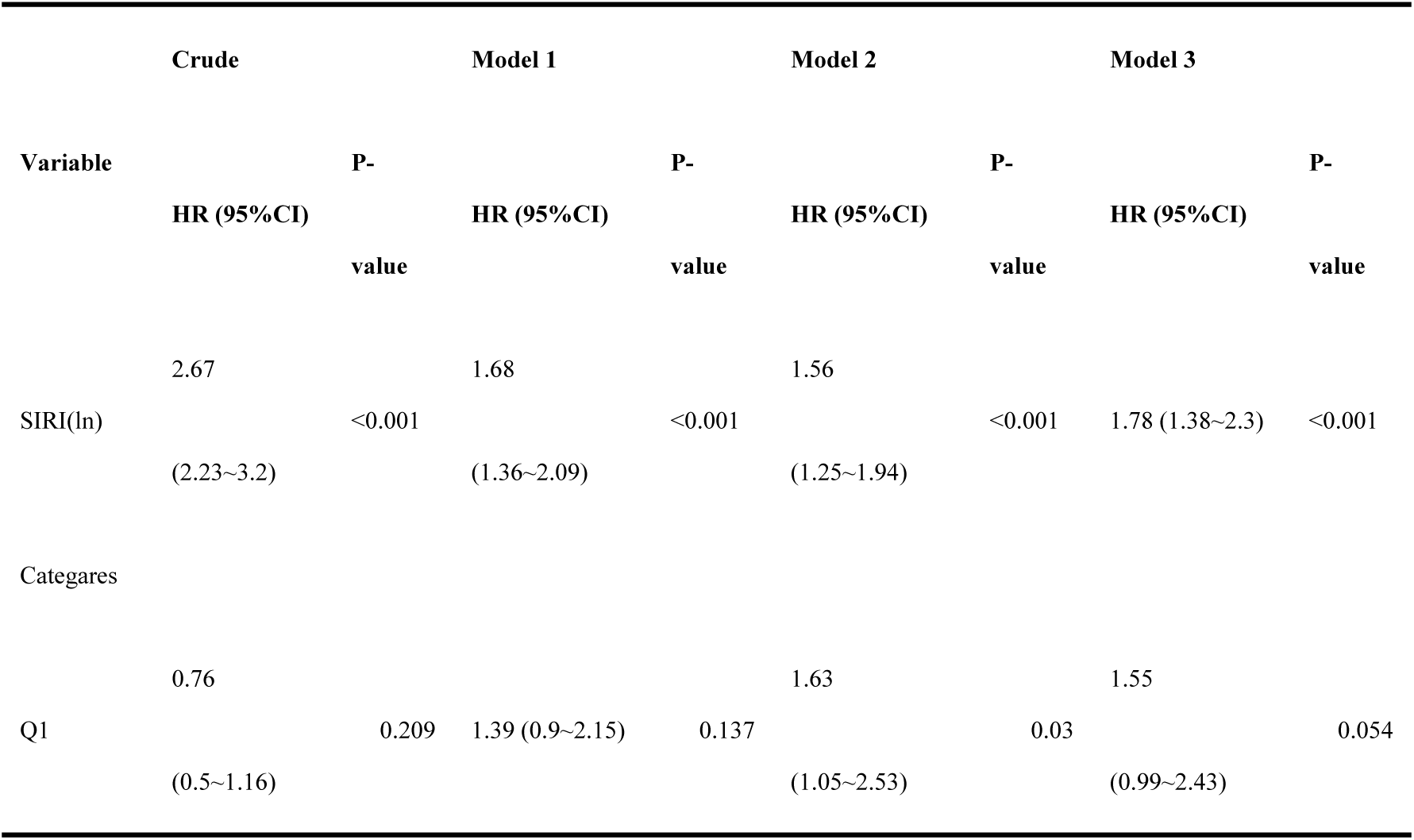
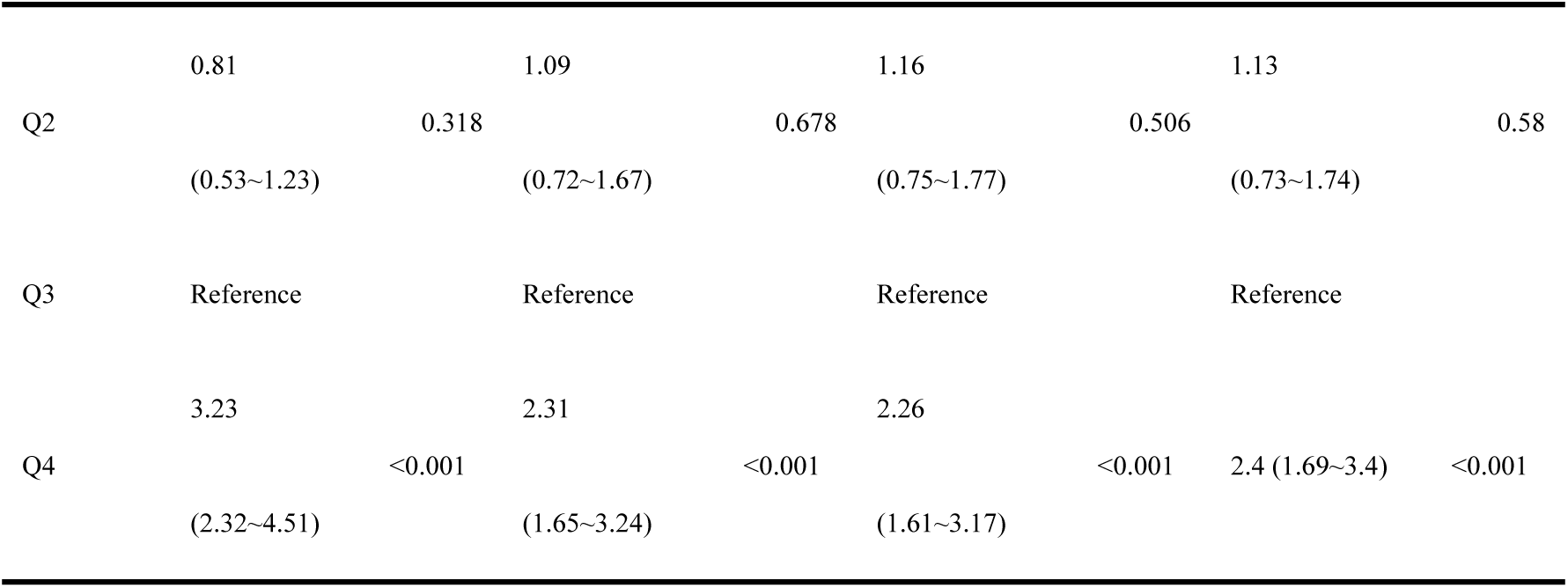
Multivariate Cox regression for SIRI on CVD mortality of participants.

In sensitivity analysis, SIRI was converted from a continuous variable to a categorical variable (Quantiles). HR, Harzard ratio; 95% CI, 95% confidence interval.

Model 1: adjusted for gender, age, race, education, marital status, income, BMI, smoking status, drinking status.

Model 2: adjusted for gender, age, race, education, marital status, income, BMI, smoking status, drinking status, Chronic medical diseases (include hypertension, coronary heart disease, stroke, diabetes mellitus, kidney disease, thyroid disease and cancer).

Model 3: adjusted for gender, age, race, education, marital status, income, BMI, smoking status, drinking status, white blood cell, Chronic medical diseases (include hypertension, coronary heart disease, stroke, diabetes mellitus, kidney disease, thyroid disease and cancer).

### Non-linear Association Between SIRI and CVD Mortality

A non-linear association was observed between SIRI and CVD mortality, with a significant threshold identified at a SIRI level of 0.244 (HR: 0.244, 95% CI: 0.217, 0.272, p < 0.001). The relationship between SIRI and CVD mortality demonstrated two distinct slopes: the first slope (below the threshold) had an HR of 0.587 (95% CI: 0.358, 0.961, p = 0.0342), and the second, steeper slope (above the threshold) had an HR of 4.807 (95% CI: 2.945, 7.845, p < 0.001) (Figure 2 and Table 4). Likelihood ratio tests confirmed that the non-linear model was significantly better than a linear model (P-value < 0.001).

**Figure 2.**
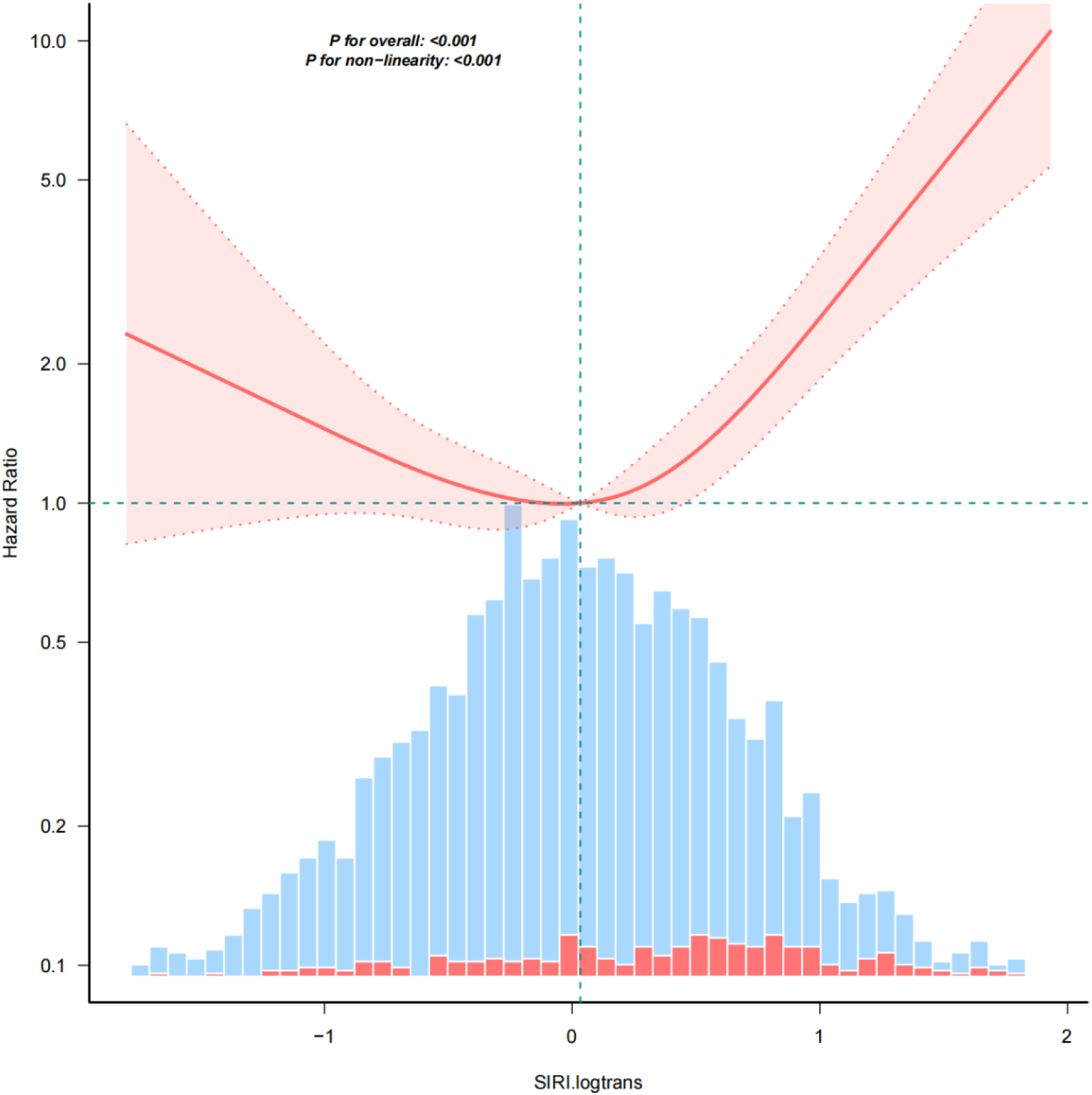
Non-linear Association Between SIRI and CVD Mortality.

**Table 4.**
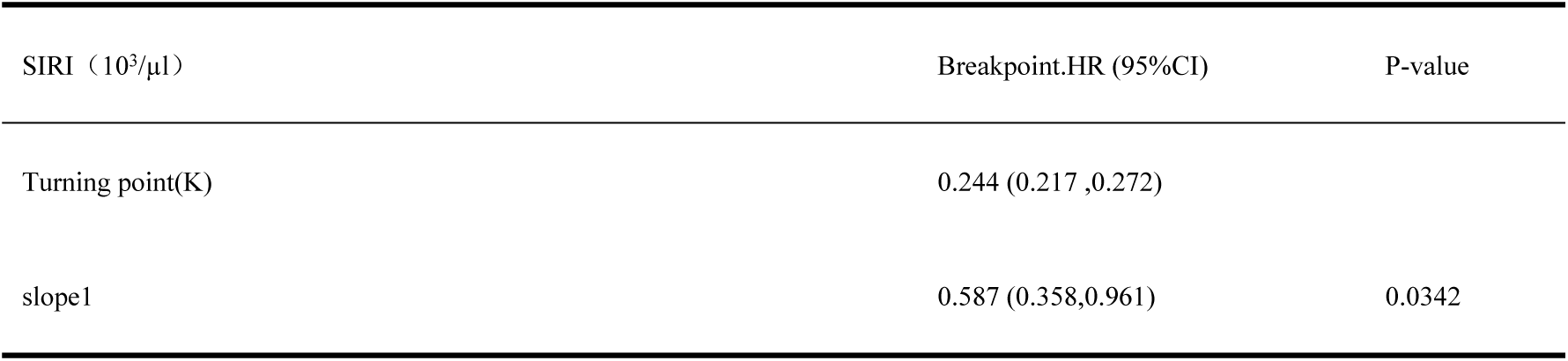
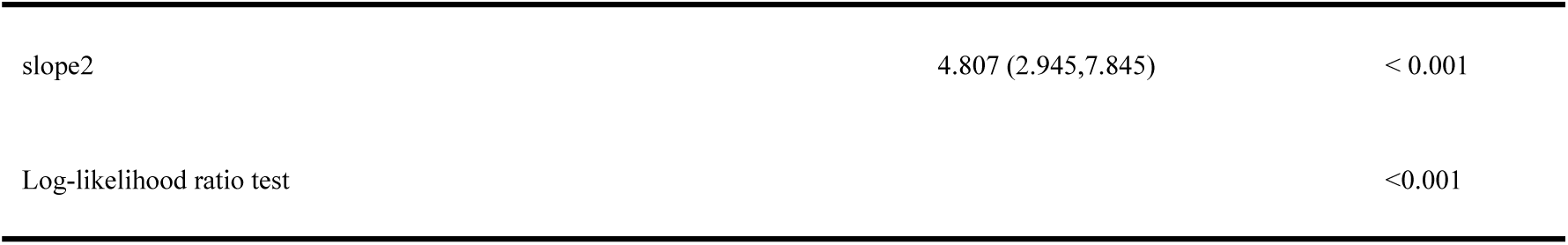
Breakpoint Analysis of SIRI and CVD Mortality.

| SIRS (10<sup>3</sup>/μl) | Breakpoint.HR (95%CI) | P-value |
| --- | --- | --- |
| Turning point(K) | 0.244 (0.217 ,0.272) |  |
| slope1 | 0.587 (0.358,0.961) | 0.0342 |
| slope2 | 4.807 (2.945,7.845) | < 0.001 |
| Log-likelihood ratio test |  | <0.001 |

### Subgroup Analysis of the Association Between SIRI and CVD Mortality

Subgroup analyses indicated that the association between SIRI and CVD mortality was generally consistent across gender, age, drinking status, BMI, CKD, diabetes, stroke, cancer, and thyroid disease (Figure 3). However, the magnitude of the association varied slightly across subgroups, with males and participants with diabetes showing higher risks. No significant interactions were found for most subgroups (all p-values for interaction > 0.05).

**Figure 3.**
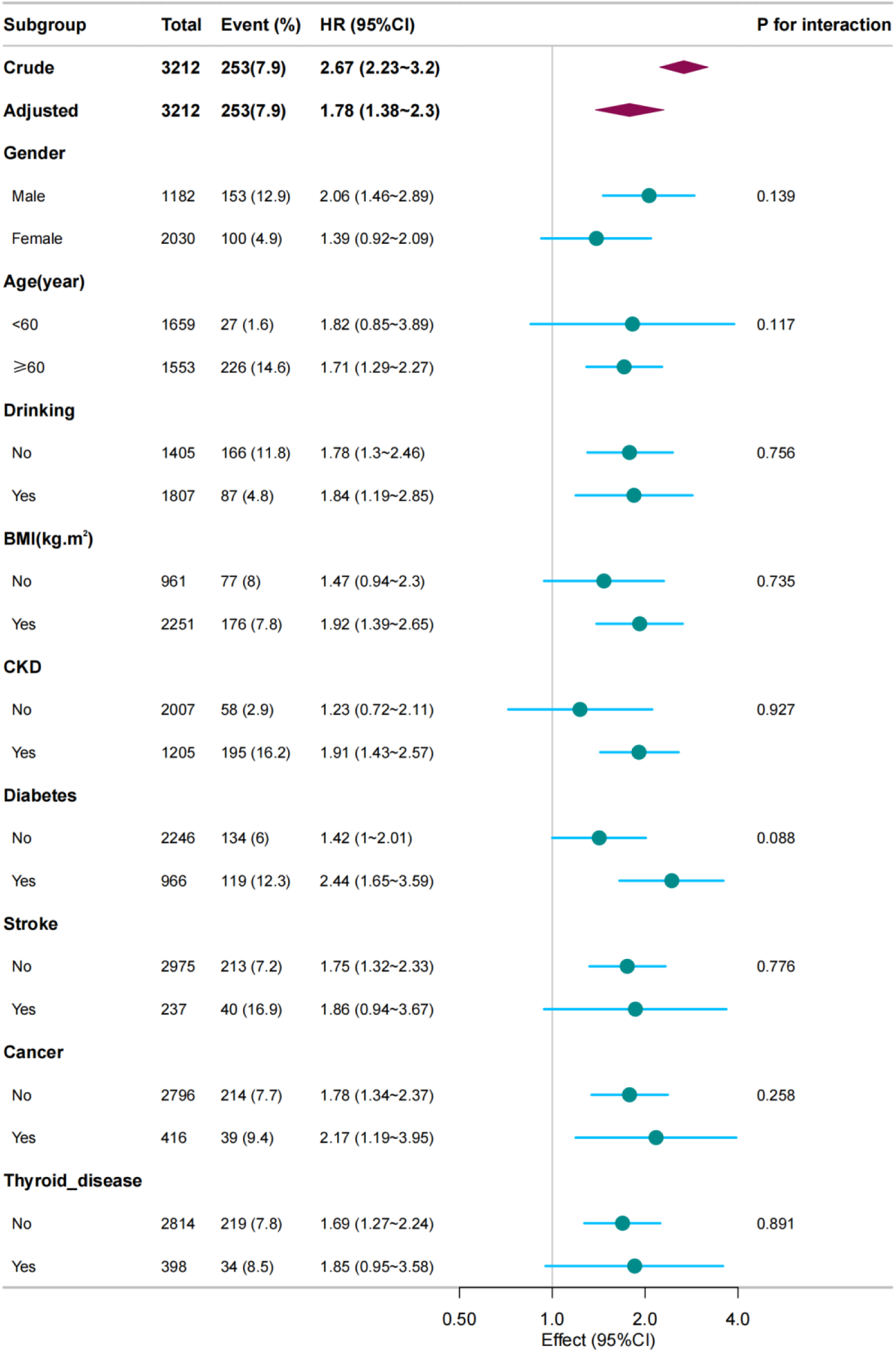
Subgroup Analysis of the Association Between SIRI and Mortality Risk.

## Discussion

This study provides compelling evidence of a significant association between the SIRI and CVD mortality in individuals with anemia. Our findings suggest that higher levels of systemic inflammation, as reflected by SIRI, are strongly linked to increased risk of CVD mortality in this vulnerable population.

Our results align with prior studies indicating that systemic inflammation plays a crucial role in cardiovascular health, especially in individuals with anemia (13, 14). The significant increase in CVD mortality with higher SIRI levels underscores the importance of inflammation in driving adverse cardiovascular outcomes. Specifically, the highest SIRI quartile exhibited a markedly elevated risk of CVD mortality compared to the lowest quartile, with hazard ratios consistently significant across both crude and adjusted models. These findings are consistent with the growing body of literature suggesting that systemic inflammation is a key factor contributing to CVD progression, particularly in populations already at higher risk, such as those with anemia.

The non-linear relationship (15) between SIRI and CVD mortality further emphasizes that inflammation may not have a linear effect on cardiovascular risk. We identified a threshold at SIRI = 0.244, above which the risk of CVD mortality steeply increased. This threshold could represent a critical level of systemic inflammation, beyond which the risk of adverse cardiovascular outcomes escalates rapidly. These findings suggest that early identification and management of inflammation, particularly in anemic individuals, may have important implications for preventing CVD mortality.

Our results have important clinical implications for the management of anemia and its associated cardiovascular risks. SIRI, as a simple and accessible biomarker, could be used to identify individuals with anemia at higher risk of CVD mortality (16). Targeting systemic inflammation in these individuals through appropriate interventions—such as anti-inflammatory treatments or lifestyle modifications—may help reduce cardiovascular morbidity and mortality (17). Additionally, our findings suggest that monitoring SIRI levels could assist healthcare providers in assessing the severity of systemic inflammation and its potential impact on cardiovascular outcomes.

subgroup analyses revealed that the association between SIRI and CVD mortality was consistent across various demographic and clinical subgroups, including gender, age, and comorbidities such as CKD and diabetes. However, the magnitude of the association was slightly stronger in males and individuals with diabetes, possibly due to the higher baseline inflammatory burden in these populations (18). The lack of significant interactions in most subgroups further supports the robustness of the observed association.

However, the study also has limitations. First, the observational design precludes the establishment of causality. While the findings suggest a strong association between SIRI and CVD mortality, further experimental and longitudinal studies are needed to confirm a causal relationship. Second, the study relied on baseline SIRI measurements, which may not fully capture the dynamic nature of systemic inflammation over time. Future studies should consider repeated measurements of SIRI to better understand its temporal relationship with CVD outcomes. Third, despite adjusting for a wide range of confounders, residual confounding cannot be entirely ruled out.

## Conclusion

In conclusion, our study demonstrates that higher SIRI levels are significantly associated with increased CVD mortality in individuals with anemia, with a non-linear relationship that suggests a critical threshold for the escalation of risk. These findings underscore the importance of systemic inflammation as a key driver of cardiovascular outcomes in this population and highlight the potential utility of SIRI as a biomarker for assessing cardiovascular risk. Further research is needed to explore the mechanisms underlying this relationship and to develop targeted strategies for managing inflammation in anemic individuals at risk for CVD mortality.

## Declarations

### Ethical Approval

Ethical approval for the research protocol and associated consent documentation (Protocol #98-12) was obtained through the NHANES Institutional Review Board’s Ethics Committee.

### Authors contributions

X.C. and L.L. acquired and analyzed the data. Y.L. and Z.L. interpreted the data. X.D.,JW and Y.T. drafted the manuscript. All authors contributed to the article and approved the final submitted version.

### Data Availability

Data supporting the results of this study can be obtained from the corresponding author.

### Consent for publication

Not applicable.

### Funding

This study was supported by Suzhou Science and Education Strengthening Health Project [Grant number:QNXM2024092].

Suzhou Science and Technology Development Program (Healthcare Science and Technology Innovation) Project (SKYD2022089)

## Acknowledgements

Not applicable.

